# Using Deep learning to Predict Cardiovascular Magnetic Resonance Findings from Echocardiography Videos

**DOI:** 10.1101/2024.04.16.24305936

**Authors:** Yuki Sahashi, Milos Vukadinovic, Grant Duffy, Debiao Li, Susan Cheng, Daniel S. Berman, David Ouyang, Alan C. Kwan

## Abstract

**Background:** Echocardiography is the most common modality for assessing cardiac structure and function. While cardiac magnetic resonance (CMR) imaging is less accessible, CMR can provide unique tissue characterization including late gadolinium enhancement (LGE), T1 and T2 mapping, and extracellular volume (ECV) which are associated with tissue fibrosis, infiltration, and inflammation. While deep learning has been shown to uncover findings not recognized by clinicians, it is unknown whether CMR-based tissue characteristics can be derived from echocardiography videos using deep learning. We hypothesized that deep learning applied to echocardiography could predict CMR-based measurements.

**Methods:** In a retrospective single-center study, adult patients with CMRs and echocardiography studies within 30 days were included. A video-based convolutional neural network was trained on echocardiography videos to predict CMR-derived labels including wall motion abnormality (WMA) presence, LGE presence, and abnormal T1, T2 or ECV across echocardiography views. The model performance was evaluated in a held-out test dataset not used for training.

**Results:** The study population included 1,453 adult patients (mean age 56±18 years, 42% female) with 2,556 paired echocardiography studies occurring on average 2 days after CMR (interquartile range 2 days prior to 6 days after). The model had high predictive capability for presence of WMA (AUC 0.873 [95%CI 0.816-0.922]), however, the model was unable to reliably detect the presence of LGE (AUC 0.699 [0.613-0.780]), native T1 (AUC 0.614 [0.500-0.715]), T2 0.553 [0.420-0.692], or ECV 0.564 [0.455-0.691]).

**Conclusions:** Deep learning applied to echocardiography accurately identified CMR-based WMA, but was unable to predict tissue characteristics, suggesting that signal for these tissue characteristics may not be present within ultrasound videos, and that the use of CMR for tissue characterization remains essential within cardiology.

**Clinical Perspective:** Tissue characterization of the heart muscle is useful for clinical diagnosis and prognosis by identifying myocardial fibrosis, inflammation, and infiltration, and can be measured using cardiac MRI. While echocardiography is highly accessible and provides excellent functional information, its ability to provide tissue characterization information is limited at this time. Our study using a deep learning approach to predict cardiac MRI-based tissue characteristics from echocardiography showed limited ability to do so, suggesting that alternative approaches, including non-deep learning methods should be considered in future research.

**Graphical Abstract:** 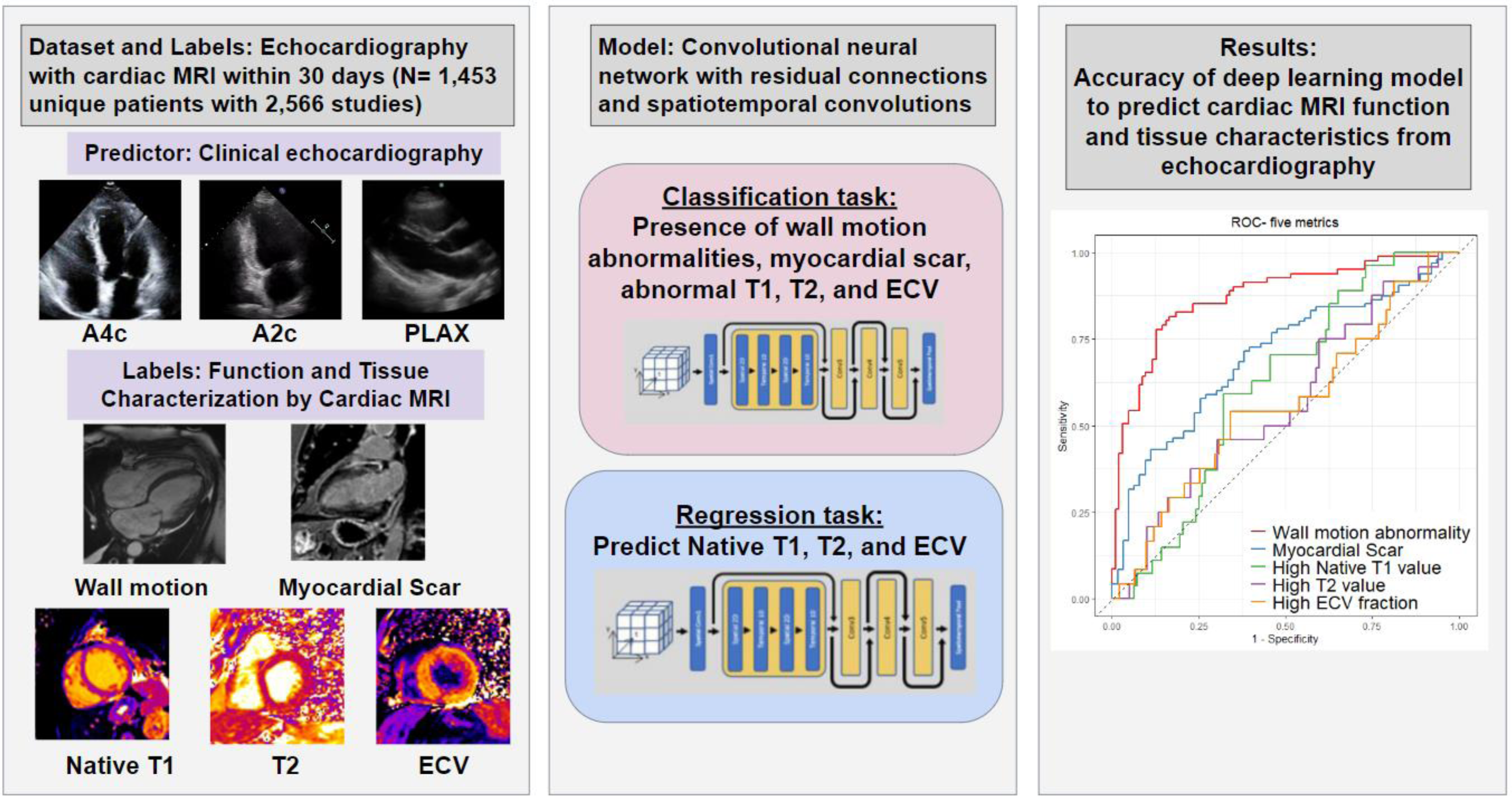

Overview of the study pipeline and results. A large echocardiography dataset involving 2,566 studies from 1,453 patients paired with CMR and echocardiography within 30 days from Cedars-Sinai Medical Center was identified. A convolutional neural network with residual connections and spatiotemporal convolutions was trained to predict each CMR finding and detect abnormal findings from echocardiography. Results showed strong prediction of functional abnormalities, but poor prediction of CMR-specific tissue characterization.

## Introduction

Echocardiography plays a central role within cardiovascular care, providing essential information on cardiac structure and function in a highly-accessible format^1^ ^2^. Cardiac MRI (CMR) is also critical to cardiovascular care but has reduced accessibility due to the availability of scanners and qualified physicians to interpret the images. In addition to providing information on cardiac structure and function, CMR is able to provide unique tissue characterization that is helpful for assessing for etiology of disease^3^. This includes myocardial composition including scarring through late gadolinium enhancement (LGE) and infiltration, edema, and diffuse fibrosis through relaxometry techniques such as T1 and T2 mapping, and extracellular volume (ECV) fraction. Despite the utility of CMR^4^, logistical barriers continue to limit its broader uptake, including the availability of CMR resources, the examination cost, the use of gadolinium contrast, and logistical complexity.

Deep learning applied to medical imaging provides the opportunity to obtain more information than currently recognized by clinicians in standard clinical care^5–9^. For example, deep learning applied to echocardiography has been shown to identify hidden features invisible to the human eye such as age, gender, serological biomarkers, prognosis, and tissue characteristics of cardiac amyloidosis and hypertrophic cardiomyopathy^10,11^. We hypothesized that deep learning applied to echocardiography could predict CMR-based measurements. In this study, we train a video-based convolutional neural network to predict CMR features from patients with paired CMR and echocardiography studies. Predicted CMR characteristics including wall motion abnormalities, myocardial scar, and markers of tissue infiltration, edema, and diffuse fibrosis were assessed for deep learning evaluation.

## Methods

### Data and study population

We identified all adults aged over 18 years at a large cardiac quaternary care center who received clinical CMR, with at least one clinical transthoracic echocardiogram within 30 days of the CMR between May 2011 and June 2022. All echocardiography were performed using Philips EPIQ 7 or iE33 ultrasound machines. Echocardiography views including apical four-chamber (A4c), apical two-chamber (A2c), and parasternal long axis (PLAX), were automatically extracted using an automated view classifier. Videos underwent automated image preprocessing including removing identifying information, electrocardiogram and respirometer tracings, and cropping and downsampling images using cubic interpolation to a standard size and resolution of 112 × 112-pixels^12^. This study was approved by the Institutional Review Board at Cedars-Sinai Medical Center and informed consent was waived due to the retrospective analysis.

Echocardiography studies were paired with the nearest CMR study within 30 days. Deep learning models were trained on echocardiogram videos with labels derived from the clincal report of the temporally closest CMR if a single echocardiography study had multiple CMRs within 30 days. Labels included presence or absence of wall motion abnormalities within the given echocardiography view (e.g., within AHA segments included within an apical 4-chamber view versus 2-chamber versus parasternal long axis) or globally (within any segment), presence or absence of LGE within a given view or globally, and both continuous and dichotomized measures of T1, T2, and ECV. Dichotomization was based on abnormal values of native T1 times over 1060ms, T2 times over 58ms, and ECV values above 33%, appropriate to scanner vendor and strength and based on local practice and published values^13^ ^14^. Overall study pipeline is demonstrated in the **Graphical Abstract**.

### CMR protocol and assessment

All CMR examinations were clinically-ordered studies performed using a 1.5T Avanto scanner (Siemens Healthineers, Erlangen, Germany). While clinical protocols varied over time, cine SSFP images were graded at the time of acquisition for the presence or absence of regional wall motion abnormalities based on an American Heart Association (AHA) 17 segmentation model ^15^. If available, T1 mapping was performed using a standard 5(3)3 modified inversion look-locker (MOLLI) sequence, with measurement of the T1 value within the mid-sepum within the mid slice. If overt tissue abnormalities were present in this region, measurement representative of the diffuse tissue composition would be performed in a secondary region, most commonly basal mid-septal slice, consistent with guidelines. If available, T2 mapping was performed using a T2-prepped SSFP sequence, with similar measurement approach as used for T1 values. Post-contrast images included T1-mapping using a short-T1 optimized MOLLI approach. ECV values were calculated from the pre- and post-contrast T1 maps in combination with point-of-care hematocrit masurement. LGE was measured 12-20 min after gadolinium contrast injection (Gadbutrol), using turbo FLASH or magnitude weighted and phase-sensitive inversion recovery gradient echo high-resolution short axis stacks, correlated with long axis LGE images, to grade presence, severity, and location of scarring using the AHA 17-segment model. All clinical examinations were reviewed by two clinicians including an advanced cardiac imaging fellow, and advanced cardiac imaging attending with Level-3 equivalent training.

### Overview of AI model and training

For model training and testing, we used a convolutional neural network with residual connections and spatiotemporal convolutions^12^ to predict CMR findings, including the presence of wall motion abnormalities, the presence of myocardial scar, native T1 value, T2 value, and ECV fraction. For binary classification tasks for predicting dichotomized CMR findings, we used binary cross-entropy loss and trained to maximize the area under the receiver operating characteristics using an AdamW optimizer with an initial learning rate of 0.001. For classification tasks including wall motion abnormalities, scarring, abnormal T1, T2, and ECV, predictions were organized by presence or absence of abnormality within the AHA segments corresponding to the specific echocardiography view. To assess for the global presence or absence of any wall motion abnormalities or myocardial scar, we combined the predictions for A4c, A2c, and PLAX views through logistic regression for a final prediction of these measurements. For a regression tasks applied for prediction of continuous labels (native T1 value, T2 value, and ECV fraction), the model was similarly trained in A4c, A2c, and PLAX views and combined to provide a global result. The model was specified to minimize the mean average error using squared loss. In both classification and regression tasks, early stopping with 10 epochs was applied, and the batch size and the number of epochs were set to 10 and 50, respectively. The dataset was randomly split at an 8:1:1 ratio for model training, validation, and held-out testing. The weights from the epoch with the best metrics were used on the held-out test dataset. All model training and evaluation were conducted using Python 3.8 and the publicly available PyTorch library.

### Statistical analysis

All performance analyses were performed using a held-out test dataset not involved in model training. For dichotomous outcomes, the model’s ability was assessed by calculating the area under the receiver operating characteristic (AUROC) curve. For continuous values including T1 value, T2 value, and ECV, mean absolute error (MAE) and coefficient of determination (R^2^) were calculated. Bland-Altman plots where the average of two measurements was plotted against the difference were used to check the agreement between the actual and predicted values from echocardiography. 95% confidence intervals were calculated with 10,000 bootstrapping samples. All data were analyzed using Python and R.

## RESULTS

### Patient characteristics

We trained and tested a model using a dataset that included 2,556 echocardiography studies paired with CMR findings from a total of 1,453 patients (mean age: 56.0±17.9 years, 41.8% female). The patient population had a range of cardiovascular comorbidities including hypertension (37.7%), hyperlipidemia (28.5%), and diabetes (16.5%) (**Table 1**). The mean left ventricular ejection fraction (LVEF) reported by echocardiography and CMR was 48.0±18.5% and 49.2±17.4%, respectively. In the CMR assessment, 48.3% had wall motion abnormalities and 49.0% had scar findings in one or more of the AHA 17 segments. Mean native T1 value was 1020±72.1 ms with 26.0% of the patients having elevated values, mean T2 value was 48.7±6.1ms with 8.2% having elevated values, and mean ECV was 28.4±5.7% with 21.8% having elevated values. The median time interval between the echocardiography and CMR was 2 days (interquartile range, −2 to +6 days).

**Table 1:**
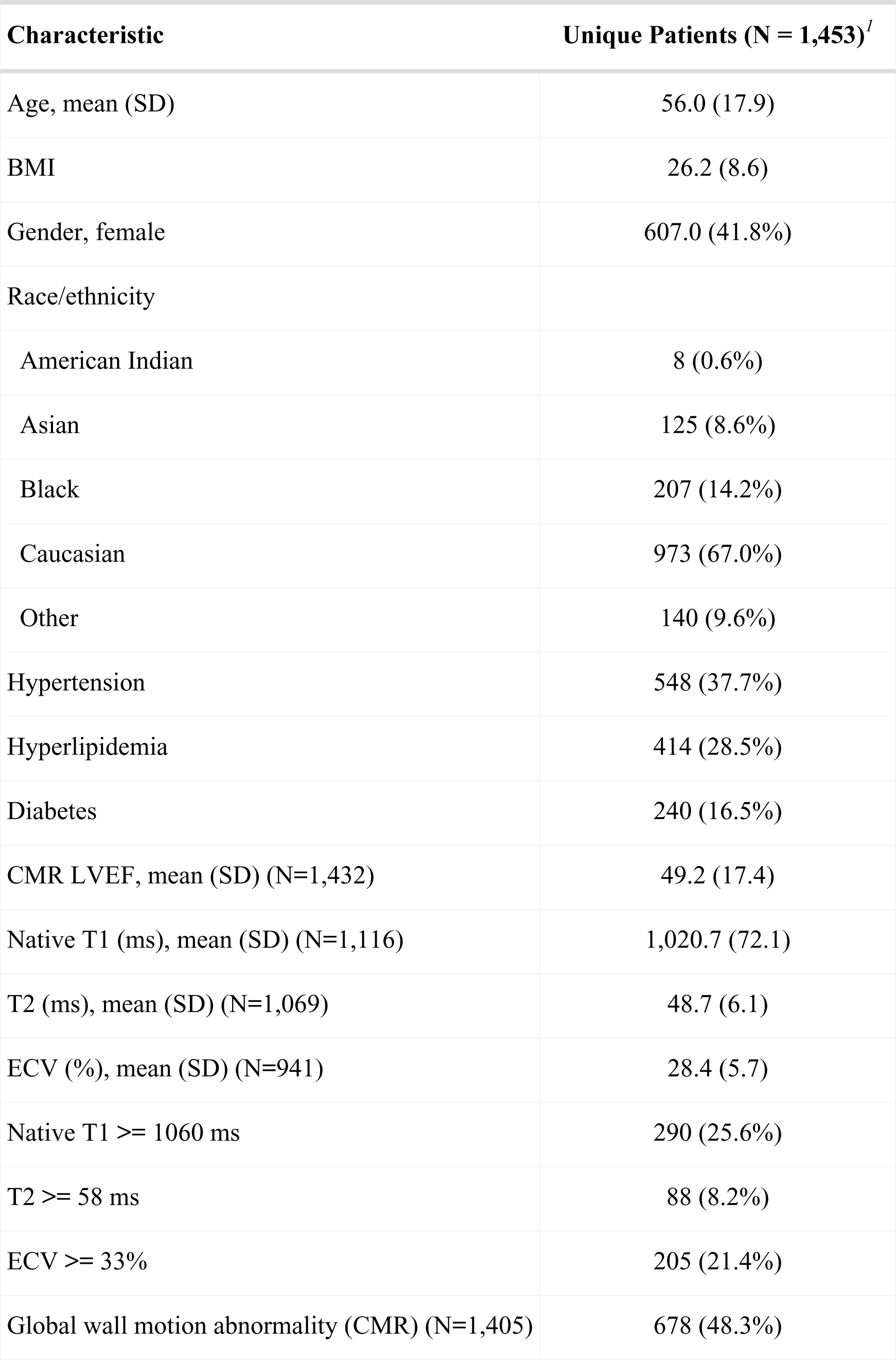
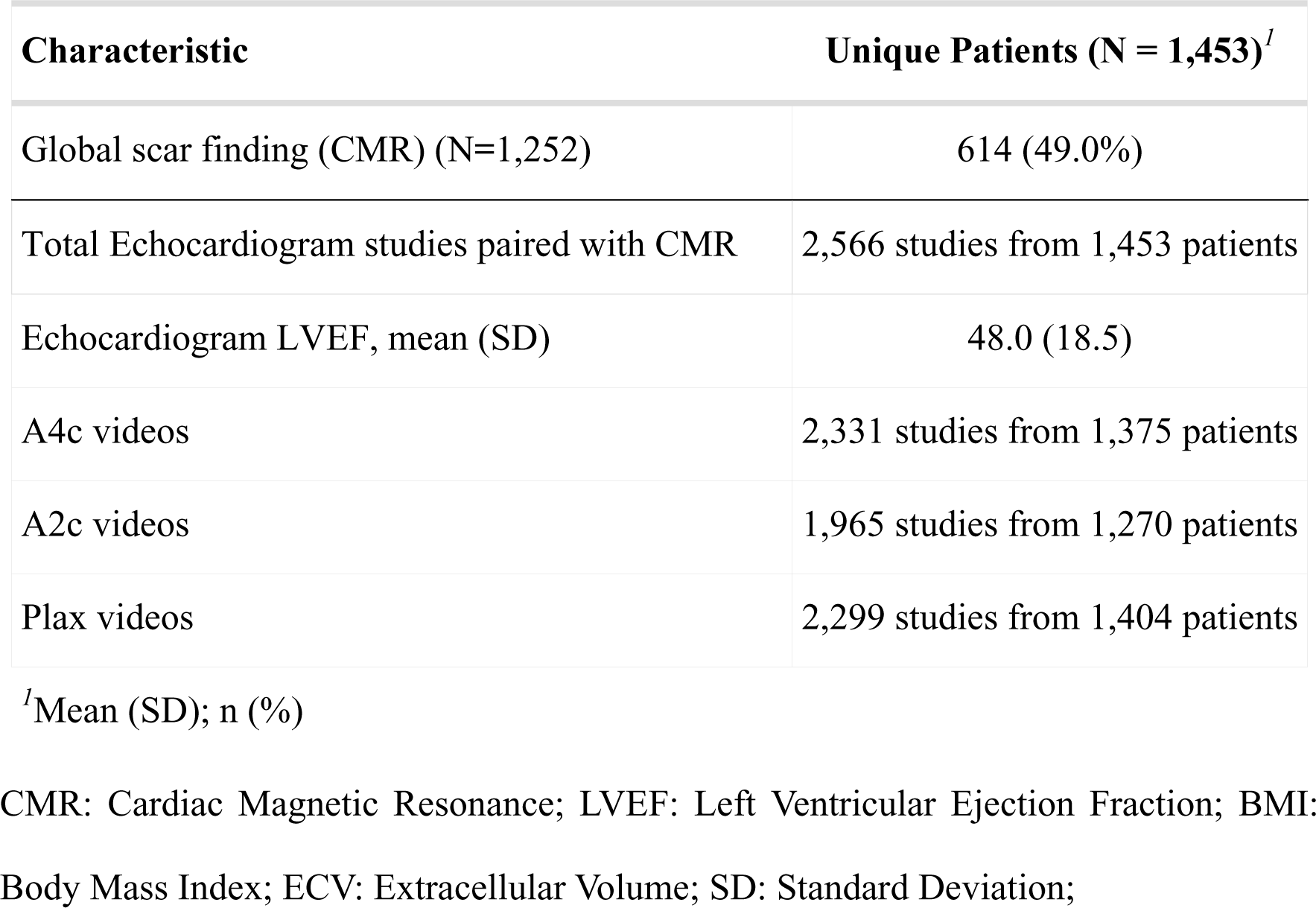
Baseline patient characteristics, cardiac magnetic resonance and echocardiogram findings.

| Characteristic | Unique Patients (N = 1,453)<sup>f</sup> |
| --- | --- |
| Age, mean (SD) | 56.0 (17.9) |
| BMI | 26.2 (8.6) |
| Gender, female | 607.0 (41.8%) |
| Race/ethnicity |  |
| American Indian | 8 (0.6%) |
| Asian | 125 (8.6%) |
| Black | 207 (14.2%) |
| Caucasian | 973 (67.0%) |
| Other | 140 (9.6%) |
| Hypertension | 548 (37.7%) |
| Hyperlipidemia | 414 (28.5%) |
| Diabetes | 240 (16.5%) |
| CMR LVEF, mean (SD) (N=1,432) | 49.2 (17.4) |
| Native T1 (ms), mean (SD) (N=1,116) | 1,020.7 (72.1) |
| T2 (ms), mean (SD) (N=1,069) | 48.7 (6.1) |
| ECV (%), mean (SD) (N=941) | 28.4 (5.7) |
| Native T1 ≥ 1060 ms | 290 (25.6%) |
| T2 ≥ 58 ms | 88 (8.2%) |
| ECV ≥ 33% | 205 (21.4%) |
| Global wall motion abnormality (CMR) (N=1,405) | 678 (48.3%) |

| Characteristic | Unique Patients (N = 1,453)<sup>1</sup> |
| --- | --- |
| Global scar finding (CMR) (N=1,252) | 614 (49.0%) |
| Total Echocardiogram studies paired with CMR | 2,566 studies from 1,453 patients |
| Echocardiogram LVEF, mean (SD) | 48.0 (18.5) |
| A4c videos | 2,331 studies from 1,375 patients |
| A2c videos | 1,965 studies from 1,270 patients |
| Plax videos | 2,299 studies from 1,404 patients |

<sup>1</sup>Mean (SD); n (%)
CMR: Cardiac Magnetic Resonance; LVEF: Left Ventricular Ejection Fraction; BMI:
Body Mass Index; ECV: Extracellular Volume; SD: Standard Deviation;

### Model Performance

Prediction of wall motion abnormalities was robust, with the AUROC for prediction within the A4c segments of 0.817 (95% CI: 0.791-0.843), A2c of 0.756 (0.707-0.802) and PLAX of 0.812 (0.777-0.847). The combination of these view prediction for global wall motion abnormalities showed strong prediction, with AUROC of 0.873 (0.816-0.922) (**Figure 1**). On the other hand, prediction of tissue composition performed poorly overall. LGE prediction was low, at AUROC of 0.657 (0.620-0.693) in the A4c views and poor at AUROC of 0.591 (0.522-0.650) in the A2c view and 0.541 (0.483 - 0.594) in the PLAX views. The global prediction of 0.699 (0.613-0.780) was the highest (**Figure 1**). AUROC for prediction of T1, T2, and ECV was similarly limited, with global prediction of abnormal T1 time of 0.614 (0.500-0.715), T2 time of 0.553 (0.420-0.692), and ECV of 0.564 (0.455-0.691). These limited capabilities were consistent across A4c, A2c, and PLAX view videos (**Table 2**). Prediction of continuous measures globally showed that the models were minimally predictive, with R^2^ of 0.04 and MAE of 49.8 ms (47.5-52.2) for T1, R^2^ of 0.002 and MAE of 5.39 ms (5.07-5.71) for T2 and R^2^ of 0.07 and MAE of 4.38% (4.17-4.58) for ECV (**Supplemental Figure 1**).

**Figure 1.**
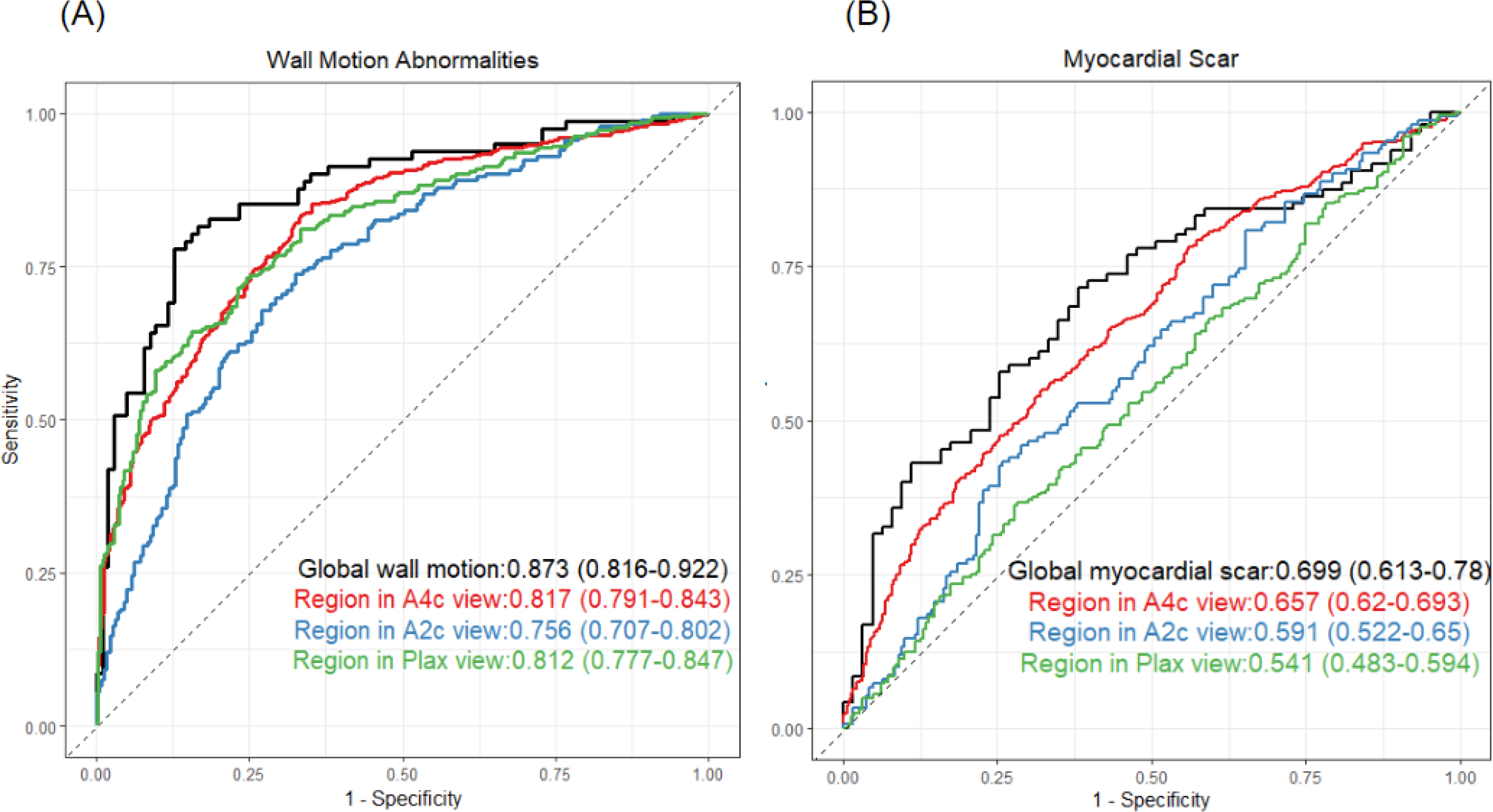
Performance of deep-learning on a held-out test dataset. Receiver-operating-characteristic (ROC) curve for predicting myocardial wall motion abnormalities and myocardial scar finding detected by CMR. A: prediction of wall motion abnormalities, B: prediction of myocardial scar. Black curves denote the performance characteristics of a deep learning model for presence of global abnormal findings. Red, blue, and green curves demonstrated the prediction of abnormal findings within A4c, A2c, and PLAX views respectively.

**Table 2:** Diagnostic accuracy of deep learning for predicting wall motion abnormalities, myocardial scar, and abnormal tissue composition by echocardiographic view.

| **Cardiac magnetic resonance imaging parameters** | **Video** | **AUROC** |
| --- | --- | --- |
| Wall motion abnormalities |  |  |
| WMA in A4c region | A4c view | 0.817 (0.791 – 0.843) |
| WMA in A2c region | A2c view | 0.756 (0.707 – 0.802) |
| WMA in PLAX region | PLAX view | 0.812 ( 0.777 – 0.847) |
|  | Logistic regression using three views | 0.873 (0.816 – 0.922) |
| Myocardial scar |  |  |
| Scar in A4c region | A4c view | 0.657 (0.620 – 0.693) |
| Scar in A2c region | A2c view | 0.591 (0.522 – 0.650) |
| Scar in PLAX region | PLAX view | 0.541 (0.483 – 0.594) |
|  | Logistic regression using three views | 0.699 (0.613 – 0.780) |
| Relaxometry parameters |  |  |
| Native T1 value (ms) | A4c view | 0.546 (0.497 – 0.596) |
|  | A2c view | 0.525 (0.479 – 0.581) |
|  | PLAX view | 0.518 (0.464 – 0.577) |
|  | Logistic regression using three views | 0.614 (0.500 - 0.715) |
| T2 value (ms) | A4c view | 0.521 (0.465 – 0.578) |
|  | A2c view | 0.539 (0.469 – 0.619) |
|  | PLAX view | 0.559 (0.491 – 0.692) |
|  | Logistic regression using three views | 0.553 (0.420-0.692) |
| ECV fraction (%) | A4c view | 0.549 (0.501 – 0.599) |
|  | A2c view | 0.598 (0.514 – 0.680) |
|  | PLAX view | 0.535 (0.463 – 0.691) |
|  | Logistic regression using three views | 0.564 (0.455 - 0.691) |

## Discussion

In this study, we investigated the accuracy of a video-based deep learning model for predicting CMR findings from echocardiography, with the express goal to bridge the gap between the accessibility of echocardiography and the diagnostic information of CMR. While the model was able to predict wall motion abnormalities, it was unable to reliably predict fundamental CMR tissue characteristics including LGE, T1, T2, and ECV. Of these, the LGE prediction was the highest at 0.699 (0.613-0.780), which is unlikely to be accurate enough for clinical utility. Overall, we would consider the attempt to predict meaningful CMR tissue characteristics from echocardiography unsuccessful.

There has been limited previous cross-modality research specifically linking echocardiography to CMR findings, with our literature review revealing no deep-learning-based publications for LGE, let alone T1, T2, and ECV. A smaller study leveraging a radiomics-based approach in patients admitted for heart failure, was able to identify the presence or absence of LGE in the anteroseptal and posterior wall myocardial segments, within the specific regions where regions of interest were placed for feature extraction. This was performed as a subset of a larger study, focusing on 89 patients for training and 40 patients for testing, all with echocardiography within 48 hours of clinical CMR, resulting in an AUROC of 0.84^16^. Other approaches such as echocardiography speckle-tracking strain has also been used to predict LGE with variable success – One publication demonstrated a significant association in 155 patients specifically diagnosed with carbon monoxide poisoning^17^, whereas a more general and smaller 50-patient population was unable to find an association with echo-based strain (AUROC of 0.58), but stronger associations with CMR-based strain (AUROC 0.67-0.78 including longitudinal and circumferential strain)^18^. Electrocardiogram (ECG) deep learning approaches have been applied to predict LGE and appear to have potentially stronger associations than were found within our study, though these are typically limited to specific populations. For example, prediction of LGE in patients with mitral valve prolapse using a CNN-based approach was able to achieve an AUROC of 0.75 in approximately 600 patients^19^, and 0.76 in a hypertrophic cardiomyopathy population of 1,930 patients though the AUROC decreased to 0.68 in external validation^20^. A smaller study in 114 patients achieved AUROC up to 0.81 for ECG prediction; however, this was from a 6-fold cross-validation without a hold-out dataset, so there is likely a significant contribution of overfitting to the model^21^.

In reconciling our results with the established literature, we note that our population was typically both larger, and more general than previous works. Inclusion of diverse clinical conditions may have reduced the ability to predict CMR tissue characteristics, as the histological correlates of LGE, T1, T2, and ECV can vary between disease processes. However, our overarching motivation for echocardiographic tissue characterization was broad accessibility independent of specific disease processes, and therefore we felt that that this was the most appropriate approach. Additionally, given that clinical echocardiography is already commonly used to accurately identify WMA, we present the strong deep learning prediction of WMA not as a proposed clinical application, but to provide quality assurance for the workflow in mapping CMR labels and echo images, and our ability to train deep learning models for echo. We also recognize that of all of the tissue characterization markers, LGE was the highest and showed small signal for prediction (AUROC of 0.699); however, without a much stronger signal, we felt this was most likely due to prediction of confounding factors such as reduced ejection fraction or thinned myocardium. These factors may be easily visible on echocardiography and are associated with, but not equivalent to LGE; and thus not clinically useful to predict with deep learning.

Overall, the results of our study suggest that at least within the current population and deep learning architecture, CMR-based tissue characteristics are unable to be derived from standard clinical echocardiography at this time. Additional experiments testing prediction of deep learning across various sample size and model architectures had only modest differences in performance (not shown). At the inception of this study, we recognized that the ability to derive magnetic resonance-specific findings from an ultrasound-based modality may have limited biological plausibility, as the image acquisition and reconstruction process between the two modalities are extremely technically distinct. Historically however, echocardiography tissue characteristics such as granular sparkling has been seen as suggestive of cardiac amyloidosis^22^ and integrated backscatter has been proposed for use within both inflammatory and fibrotic conditions^23,24^. Detection of histological characteristics by echocardiography thus may be best directly quantified, as while CMR-based tissue characterization is well-accepted, it still represents a surrogate of the true tissue composition. Novel echocardiography techniques such as shear-wave elastography can provide signal for fibrosis not available through standard clinical images and may be able to expand the applicability of echocardiography independent of deep-learning based techniques^25,26^. Thus, while our results support the ongoing utility for CMR independent of echocardiography, we are optimistic for the future of echocardiography to provide highly accessible tissue characterization.

### Limitations

There are several limitations in the present study. First, as a retrospective single center study, our results were the result of a limited dataset. In particular, CMR practices may vary significantly between locations, and it remains possible stronger relationships can be found with larger datasets. The 30-day interval was selected as being a relatively short time frame but with a high number of eligible studies, and we recognize that incident clinical events or resolution of acute findings may have occurred between the two studies, though the short median time interval gives some degree of assurance. CMR referral was clinical and included a wide range of diseases conditions and severities. This heterogeneity may have increased the challenge of finding significant associations.

## Conclusion

In conclusion, we found that a video-based deep learning architecture using echocardiography was able to identify CMR-based WMA, but was unable to accurately identify CMR-based tissue characteristics including LGE, T1 time, T2 time, and ECV. Further testing using alternative populations and approaches should be considered. At present, our study supports the ongoing use of CMR for tissue characterization in appropriate patients, despite challenges to patient access.

## Sources of Funding

ACK: AHA 23CDA1053659, NIH UL1TR001881, 75N92020D00021

DO: NHLBI R00HL157421, 75N92020D00021

YS: Japanese Society for the Promotion of Science, Grants-in-Aid for Scientific Research (JSPS-KAKENHI)

## Author contributions

Concept and design: YS, DO, AK

Acquisition, analysis, or interpretation of data: YS, DO, AK, MV, GD, DB, SC

Drafting and Critical revision of the manuscript: All authors

Statistical analysis: YS, DO, AK

Obtained funding: DO, YS, AK

Supervision: AK, DO, SC, DB, DL

Data and code availability

All of the code for the present study is available at https://github.com/echonet/.

## Abbreviations

AHA: American Heart Association
AUROC: Area under receiver operating characteristic
A4C: Apical 4 chamber
A2C: Apical 2 chamber
CMR: Cardiac magnetic resonance
ECV: Extracellular volume
LVEF: Left ventricular ejection fraction
LGE: late gadolinium enhancement
PLAX: Parasternal long axis
MOLLI: modified inversion look-locker

## Data Availability

Data available on reasonable request

---

**Supplementary Figure 1:**
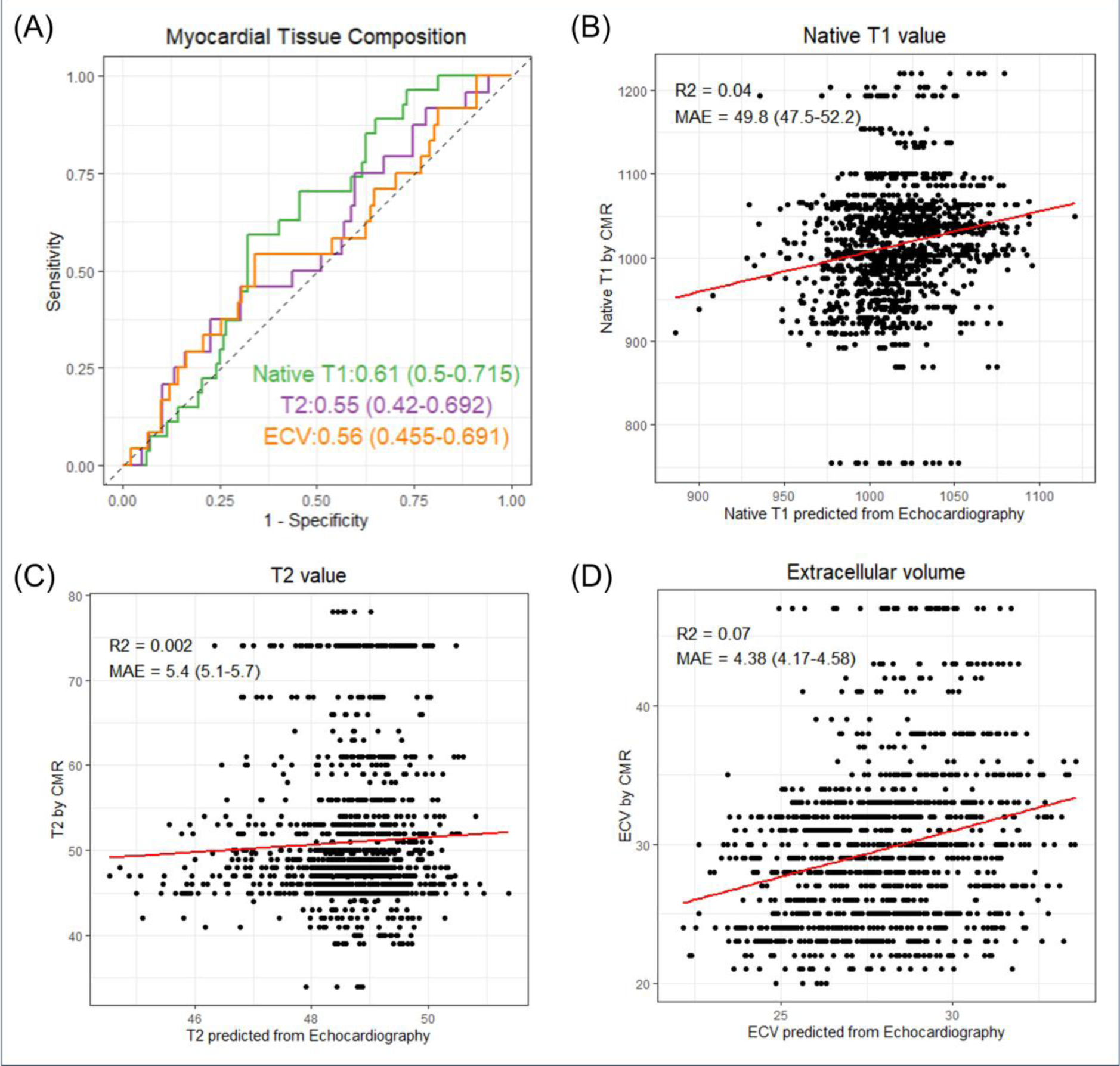
Performance of deep-learning model on prediction of abnormalities and estimation of CMR-specific tissue composition. (A) Receiver-operating-characteristic (ROC) curve for predicting abnormal CMR findings, including native T1 value, T2 value, and extracellular volume fraction. Green curve denotes the prediction of native T1 finding ≥1060ms. Purple denotes the prediction of T2 ≥ 58ms. The orange curve denotes the prediction of ECV ≥33%. Scatterplot for predicted versus measured (B) native T1 value, (C) T2 value, and (D) ECV.

